# Unraveling the Potential of Physical Activity and High-Intensity Interval Training in the Treatment of Familial Hypercholesterolemia: Impact on Cardiorespiratory Fitness, Atherosclerosis, and Underlying Mechanisms. Study protocol of the UPPA-FH Project

**DOI:** 10.64898/2025.12.22.25342692

**Authors:** José Antonio Vargas Hitos, Pablo González-Bustos, María José Martín-Jáimez, Antonio Díaz Chamorro, Blanca Sánchez Checa, Alberto Ortiz Parra, Lauro Quesada Jiménez, Pablo García-Mateo, Elena Martínez Rosales, Cristian García Sánchez, Andrés Baena-Raya, Alba Hernández-Martínez, Rocío Sánchez-Sánchez, María Emiliana Bellón Guardia, Yolanda García Rivero, Antonia Román Muñoz, Álvaro Contreras-Rodríguez, Antonio Jesús Láinez Ramos-Bossini, Genaro López-Milena, Ricardo Rivera López, Rafael Peñafiel Burkhardt, Víctor López Espinosa, María del Señor López Vélez, Fernando Rodríguez Alemán, Javier Blanco-Ramos, Ismael Francisco Aomar-Millán, Juan Antonio Montes-Romero, María Luisa Fernández-Almira, Purificación Sánchez-López, Tania Romacho, Ana Cristina Abreu, Ignacio Fernández, Fernando Jaén-Águila, Juan Diego Mediavilla-García, Nuria Navarrete-Navarrete, Blanca Gavilán-Carrera, Alberto Soriano Maldonado

## Abstract

**Introduction:** Familial hypercholesterolemia (FH) is a genetic disorder characterized by elevated low-density lipoprotein -cholesterol (LDL-c) levels, increasing early cardiovascular risk. Many patients do not reach LDL-c targets despite treatment. Physical activity and exercise may help by improving cardiorespiratory fitness (CRF), reducing inflammation, and modulating metabolic pathways. This study aims 1) to cross-sectionally evaluate the association of physical activity and CRF with markers of sub-clinical atherosclerosis and a nuclear magnetic resonance-derived metabolomic profile in patients with FH, and 2) to assess the effects of high-intensity interval training (HIIT) and moderate-intensity continuous training (MICT) on CRF (primary outcome), markers of subclinical atherosclerosis, serum biomarkers and metabolomic profiles (secondary outcomes), and to unravel the underlying mechanisms.

**Methods and analysis:** For aim 1, a cross-sectional study will be conducted in approximately 200 patients with FH from Granada and Almería (Spain). Assessments will include accelerometer-measured physical activity, CRF, markers of subclinical atherosclerosis and metabolomic profiles. For aim 2, a 16-week, parallel-group randomized controlled trial will be conducted with 75 participants assigned to one of three groups: HIIT (4 intervals of 4 minutes at 85–95% of maximal heart rate, 3 days/week), MICT (34 minutes at 69–76% of maximal heart rate, 3 days/week) or usual care. CRF will be assessed using the modified Bruce test. Markers of subclinical atherosclerosis will include vascular inflammation (PET/CT scan), arterial stiffness (Mobil-O-Graph® 24h pulse wave monitor), carotid intima-media thickness, and carotid plaque presence (carotid Doppler ultrasound). Metabolomic profiles will be analyzed using nuclear magnetic resonance spectroscopy. Analyses will include correlation and regression models for cross-sectional associations, and linear mixed-effects models for RCT outcomes, following an intention-to-treat approach with additional sensitivity analyses.

**Ethics and dissemination:** The study was approved by the Ethics Committee for Biomedical Research of Granada (ref. 1417-N-23), and findings will be disseminated through peer-reviewed publications, conference presentations, and outreach activities aimed at patients and healthcare professionals.

**Trial registration number:** ClinicalTrials.gov ID NCT06833944.

**Strengths and limitations of this study:**

- This is the first study to evaluate the effects of structured exercise in patients with familial hypercholesterolemia, comparing two exercise modalities (high-intensity interval training and moderate-intensity continuous training) and assessing multiple clinically relevant health outcomes (cardiorespiratory fitness, subclinical atherosclerosis, and metabolomic profiles).
- The study integrates objective measures of physical activity (accelerometry), cardiorespiratory fitness (VO□max), vascular imaging (PET/CT, carotid ultrasound, arterial stiffness), and metabolomics, providing a comprehensive assessment of both functional and mechanistic outcomes.
- The interventions are supervised, individualized, and follow established exercise reporting guidelines (CERT), which enhances safety, adherence, and reproducibility.
- The intervention duration (16 weeks) and the specific clinical population (aged 18– 70 years, clinically stable, without cardiovascular disease) may limit assessment of long-term effects and generalizability to all patients with familial hypercholesterolemia.
- Although some outcomes are directly applicable to clinical or exercise practice, other mechanistic outcomes (PET/CT, metabolomics, and vascular imaging) provide rigorous scientific insight but have lower direct applicability.

## INTRODUCTION

Familial hypercholesterolemia (FH) is an inherited disorder characterized by elevated low-density lipoprotein cholesterol (LDL-c) ^1^ that affects around 25 million people worldwide ^2^. Individuals with FH have a 13-fold higher risk of atherosclerotic cardiovascular disease (CVD) ^3^, with up to 50% of men and 30% of women experiencing myocardial infarction before age 60 ^4^. FH requires lifelong management, often with multiple medications, imposing a substantial burden on healthcare systems. In Spain, it generates over €87 million annually in direct and indirect costs ^5^.

FH is primarily managed with pharmacological therapies (statins, ezetimibe, and PCSK9 inhibitors), which can reduce LDL-c by up to 60% ^6^. However, only 30–40% of patients reach LDL-c targets, often due to statin intolerance ^7^, underscoring the need for complementary strategies ^8^. Detecting subclinical atherosclerosis is key, as it reflects cumulative LDL-c exposure alongside endothelial dysfunction, low-grade inflammation, and oxidative stress ^9^. In this context, positron emission tomography (PET) combined with plasma markers of inflammation and oxidative stress such as high sensitive C-reactive protein [hsCRP], oxidized LDL [oxLDL], or soluble intracellular adhesion molecule 1 [sICAM1] provides valuable information on vascular inflammation and helps predict plaque progression and future cardiovascular events ^10,11^.

Physical activity is a well-established predictor of health and survival in both the general population ^12^ and patients with CVD ^13^. Likewise, cardiorespiratory fitness (CRF) is a powerful health marker, proposed as a clinical vital sign ^14^, and is linked to healthier lipid profiles, and lower rates of hypertension, diabetes, subclinical atherosclerosis, and all-cause CVD mortality ^15^. Despite strong evidence, the cardioprotective role of physical activity and CRF is often overlooked, and their effects on key CVD mechanisms in FH (low-grade chronic inflammation, endothelial dysfunction or atherosclerosis progression) are still poorly understood.

Evidence on physical activity in FH is limited to behavioral interventions showing minimal effects on traditional CVD risk factors ^16,17^, or observational studies linking higher activity to lower CVD incidence ^18^. Although dyslipidemia guidelines recommend ≥150 min/week of moderate aerobic activity ^6,19^, this general advice rarely targets the mechanisms driving atherosclerosis in FH. Structured exercise, however, offers broader benefits: reducing inflammation and oxidative stress, improving endothelial function, providing cost-effective outcomes comparable to pharmacological therapies, and uniquely enhancing CRF ^20^. Traditionally, moderate-intensity continuous training (MICT) has been prescribed to improve cardiometabolic health, but high-intensity interval training (HIIT) may provide superior benefits for CRF ^21^ and vascular function ^22^. These effects likely arise from enhanced shear stress, nitric oxide, vasodilation, and anti-inflammatory, antioxidant, and metabolic effects, particularly in individuals with metabolic disorders or CVD ^22^. Given its safety ^23^, time efficiency, and higher enjoyment ^24^, HIIT may represent a more efficient strategy to enhance CRF and mitigate atherosclerosis in FH.

Despite compelling evidence supporting the benefits of physical activity and CRF on cardiometabolic risk, their role in key cardiometabolic outcomes in patients with FH remains unexplored. Similarly, no prior randomized trials (RCTs) have tested the effects of structured exercise on cardiometabolic risk and underlying mechanisms in this population. Therefore, the UPPA-FH study has two general aims: 1) to evaluate the association of physical activity and CRF with markers of subclinical atherosclerosis and the metabolomic profile in patients with FH (cross-sectional study), and 2) to assess the effects of HIIT and MICT compared to usual care, on CRF (primary outcome), markers of subclinical atherosclerosis, serum biomarkers and metabolomic profile (secondary outcomes), and to unravel the underlying mechanisms (RCT).

## METHODS

### Study Design

The UPPA-FH project proposes two complementary study designs. In 2024, a cross-sectional study started to address aim 1. Concurrently, a subsample meeting the inclusion criteria was recruited for a three-arm parallel group RCT to address aim 2. Interventions will be conducted in two waves, in 2025 and 2026. **Figure 1** shows a graphical representation of the study design. The study protocol was approved by the Ethics Committee for Biomedical Research of the Province of Granada (CEI/CEIM Granada ref: 1417-N-23) before recruitment. The RCT was registered at clinicalstrials.gov on January 27, 2025 (NCT06833944). The protocol follows the main principles outlined in the SPIRIT reporting guideline (Standard Protocol Items: Recommendations for Interventional Trials) ^25^. The results will be reported according to CONSORT standards (http://www.consort-statement.org/).

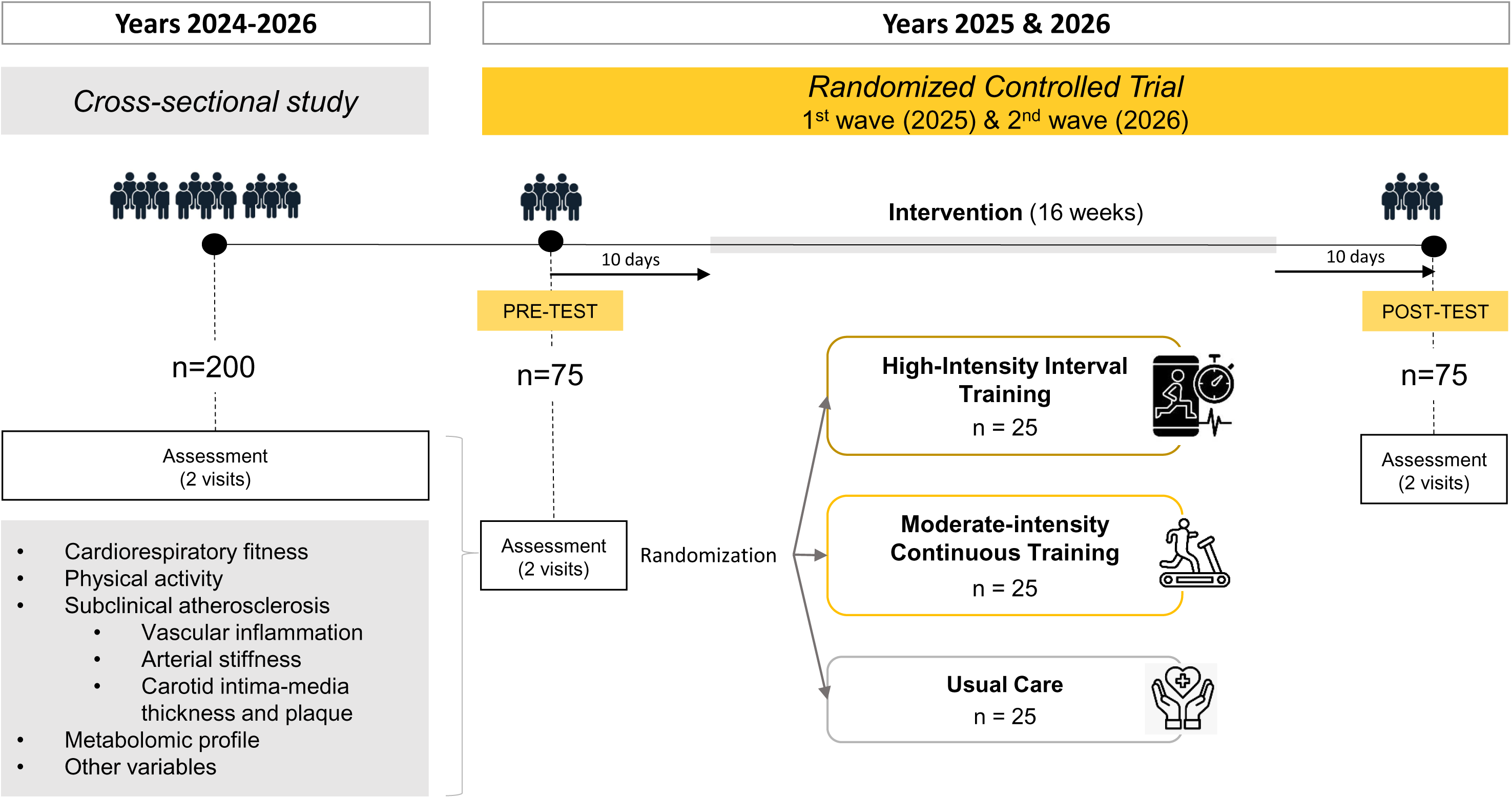

### Recruitment and Eligibility Criteria

Patient recruitment for the cross-sectional study will occur in 2024–2026. Volunteers aged 18–70 with genetically confirmed FH ^1^ and at least 1 year of follow-up at the Vascular Risk Units of the Hospital Universitario Virgen de las Nieves (HUVN), Hospital Universitario San Cecilio (HUSC) or Hospital Universitario Torrecárdenas (HUT) will be recruited. For the RCT, in addition to the inclusion criteria described for the cross-sectional study, participants aged > 45 years will also be required to demonstrate clinical and treatment stability, as well as a non-pathological coronary computed tomography (CT) angiography performed within the past 24 months. Exclusion criteria for both studies include: (i) inability to provide informed consent, (ii) exercise contraindications, and (iii) history of clinical CVD (ischemic heart disease, stroke, or peripheral arterial disease). Participants will be asked to maintain their usual diet throughout the study.

### Sample Size, Randomization, and Blinding

For the cross-sectional study, ∼200 voluntary FH patients followed at the Vascular Risk Units of HUVN, HUSC, or HUT will be recruited. This real-life cohort is expected to provide sufficient precision to examine associations between physical activity, CRF, and markers of subclinical atherosclerosis. Assuming small-to-moderate effect sizes (r≈0.20–0.30), 80% power, and α=0.05, the minimum required sample is 191 participants.

For the RCT, no prior data exist on CRF changes following exercise in FH. Based on evidence from healthy and obese adults, HIIT can induce large improvements in CRF ^26^. Considering population differences, the sample size was conservatively set to detect a medium-high effect size (f=0.40) with 80% power and a two-sided α of 0.05, yielding a minimum of 66 participants (22 per group). Accounting for a 15% anticipated dropout, 75 participants (25 per group) will be recruited. Adherence strategies will be implemented to minimize loss to follow-up (see Potential Risks, Adverse Events, and Contingency Plan).

RCT participants will be randomized 1:1:1 to HIIT, MICT, or control in a superiority framework. A sex-stratified randomization sequence will be generated by an independent researcher using a computer-based random number generator and securely stored by personnel not involved in recruitment or assessment. Assignments will be placed in sequentially numbered, opaque, sealed envelopes and opened only after participants meet all inclusion criteria, provide informed consent, and complete baseline assessments. Due to the nature of the intervention, participants cannot be blinded, but outcome assessors and data analysts will remain blinded throughout the study.

### Data Collection

#### Assessment Protocol

Cross-sectional study: The study will involve two on-site visits at HUVN or HUT. Before the first visit, sociodemographic and clinical data will be collected, and participants will complete validated online questionnaires assessing anxiety, depression, quality of life, and adherence to the Mediterranean diet. During the first visit, participants will undergo carotid Doppler ultrasound, pulse wave velocity measurement, anthropometry, and a battery of physical fitness tests. They will then receive an accelerometer to wear continuously for 10 consecutive days. During the second visit, fasting blood samples will be collected, followed by PET/CT and dual-energy X-ray absorptiometry (DXA) scans, and the accelerometer will be retrieved.

Randomized controlled trial: The same procedures and measures as in the cross-sectional study will be applied. Assessments of all study variables will be conducted at baseline (within 10 days before the intervention) and post-intervention (within 10 working days after completion). Each assessment will take place over two non-consecutive days at HUVN or HUT, with at least 10 days between them.

#### Primary Outcome

CRF: the maximum oxygen uptake (VO_2max_) will be continuously measured using the Quark cardiopulmonary exercise testing (CPET) metabolic analysis system from COSMED (reference COS-C09073-01 1-99), by performing the ramped Bruce protocol^27^. This test is performed on a treadmill with 15 seconds stages with speed and grade increasing progressively (i.e., matching the speed and grade of the original Bruce protocol at the end of each 3-min stage) (see details in online **Supplemental Table 1**). VO_2max_ will be established following ACSM standards ^28^ by identifying an oxygen uptake plateau (defined as an increase in VO_2_ of less than 150 mL/min between two successive work rates) as the sole validation criterion. If no plateau is observed, VO_2max_ will only be accepted when two or more of the following auxiliary thresholds are met: peak respiratory exchange ratio (RER) ≥ 1.10; peak heart rate (HR) ≥ 85 % of age-predicted maximum (208 – 0.7 x age) ^29^; and peak rating of perceived exertion (RPE) ≥ 7 on the Borg 0–10 scale ^30^. In cases where these criteria are not satisfied, the highest value reached will be reported as peak oxygen uptake (VO_2peak_). The RPE (1-10 Borg scale), maximal HR (HR_max_, bpm at exhaustion), recovery HR and blood pressure (1, 2, and 3 minutes later, bpm), and the use of medications potentially affecting HR (e.g., beta-blockers or digoxin) will be recorded.

#### Secondary Outcomes

The secondary outcomes, along with their assessment procedures, are presented in **Table 1**. Further details regarding these outcomes and procedures can be found in the online **Supplementary Material** (pp. 1-8).

**Table 1.**
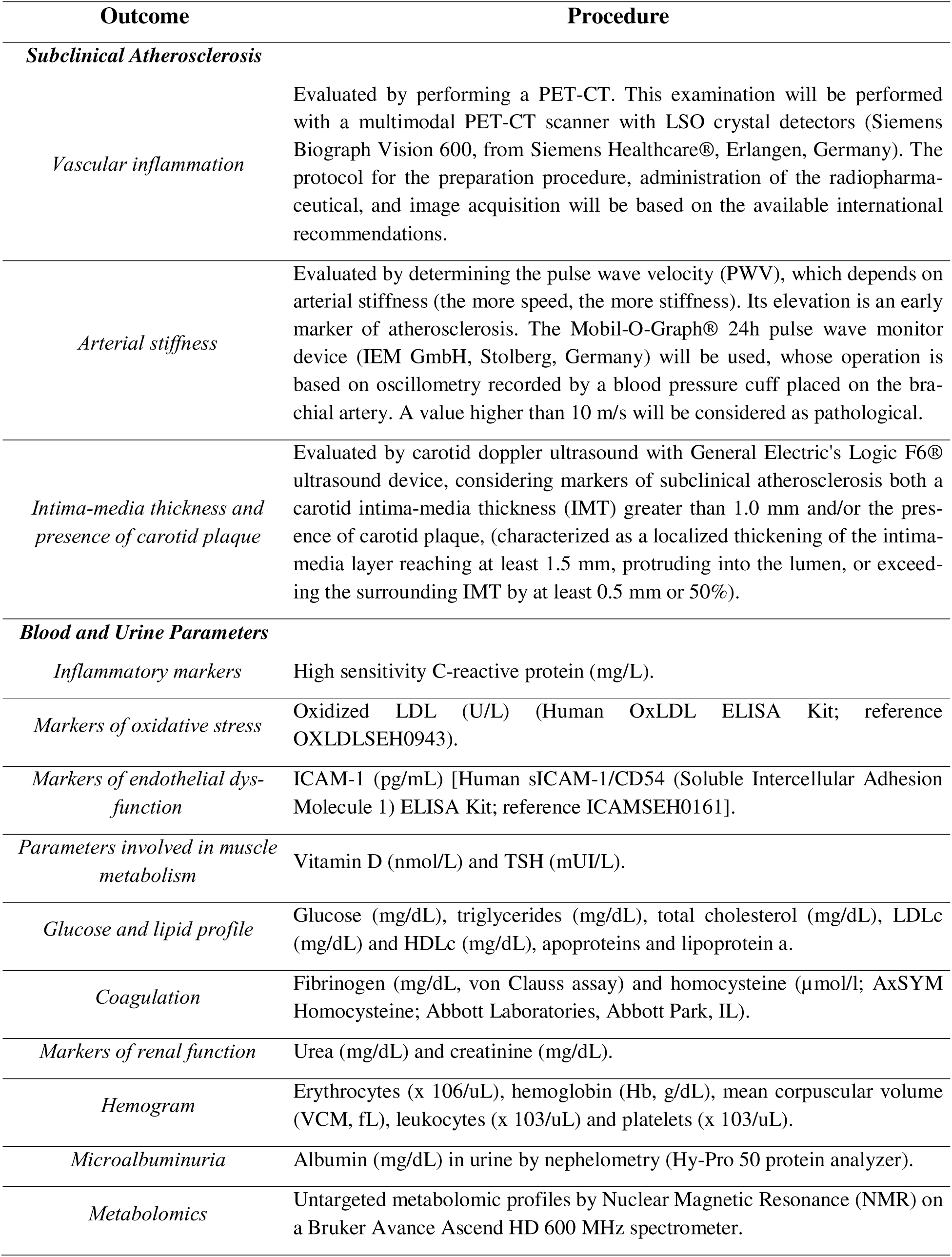
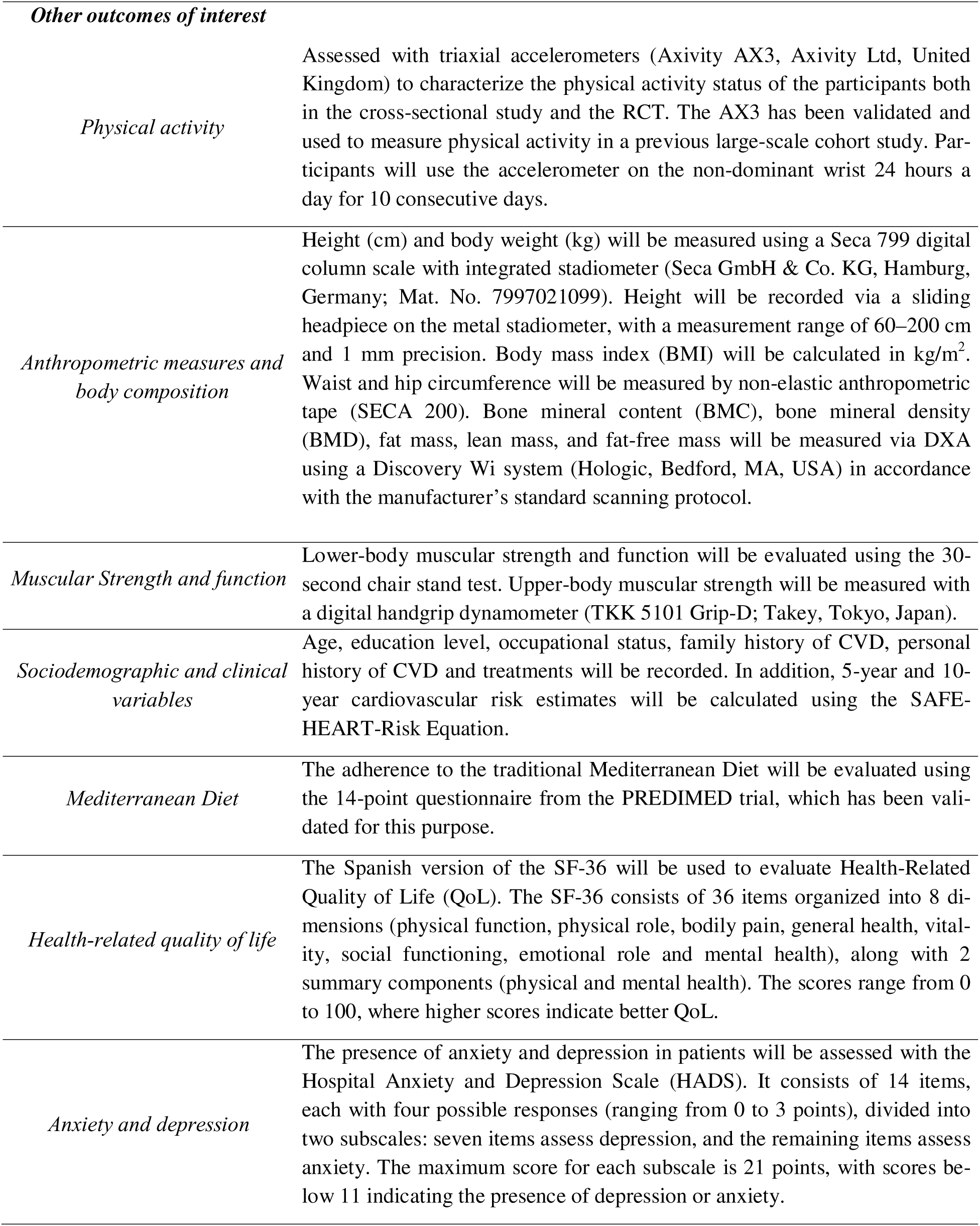
Secondary outcomes and procedures of assessment.

### Intervention

To maximize transparency and replicability, the exercise protocols (HIIT and MICT) followed the *Consensus on Exercise Reporting Template* (CERT) ^31^, which provides detailed guidelines for reporting essential parameters in exercise intervention trials (see **Table 2**). The intervention will consist of a 16-week HIIT or MICT program, or a waiting-list control group that will not perform the intervention during the study period.

**Table 2.**
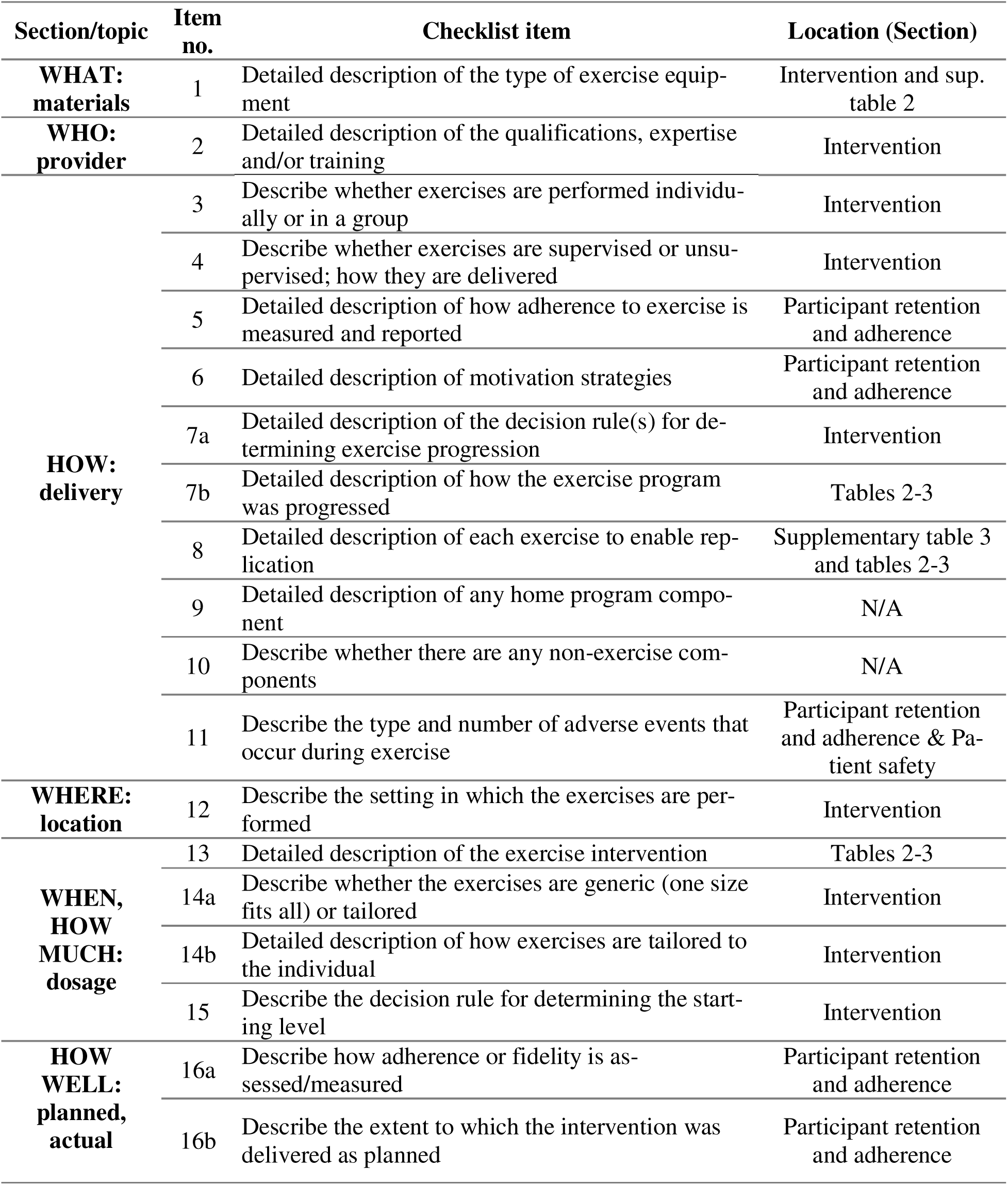
Consensus on Exercise Reporting Template (CERT) checklist from the UPPA-FH intervention trial.

| Section/topic | Item no. | Checklist item | Location (Section) |
| --- | --- | --- | --- |
| **WHAT: materials** | 1 | Detailed description of the type of exercise equipment | Intervention and sup. table 2 |
| **WHO: provider** | 2 | Detailed description of the qualifications, expertise and/or training | Intervention |
| **HOW: delivery** | 3 | Describe whether exercises are performed individually or in a group | Intervention |
|  | 4 | Describe whether exercises are supervised or unsupervised; how they are delivered | Intervention |
|  | 5 | Detailed description of how adherence to exercise is measured and reported | Participant retention and adherence |
|  | 6 | Detailed description of motivation strategies | Participant retention and adherence |
|  | 7a | Detailed description of the decision rule(s) for determining exercise progression | Intervention |
|  | 7b | Detailed description of how the exercise program was progressed | Tables 2-3 |
|  | 8 | Detailed description of each exercise to enable replication | Supplementary table 3 and tables 2-3 |
|  | 9 | Detailed description of any home program component | N/A |
|  | 10 | Describe whether there are any non-exercise components | N/A |
|  | 11 | Describe the type and number of adverse events that occur during exercise | Participant retention and adherence & Patient safety |
| **WHERE: location** | 12 | Describe the setting in which the exercises are performed | Intervention |
| **WHEN, HOW MUCH: dosage** | 13 | Detailed description of the exercise intervention | Tables 2-3 |
|  | 14a | Describe whether the exercises are generic (one size fits all) or tailored | Intervention |
|  | 14b | Detailed description of how exercises are tailored to the individual | Intervention |
|  | 15 | Describe the decision rule for determining the starting level | Intervention |
| **HOW WELL: planned, actual** | 16a | Describe how adherence or fidelity is assessed/measured | Participant retention and adherence |
|  | 16b | Describe the extent to which the intervention was delivered as planned | Participant retention and adherence |

#### Group 1: HIIT intervention program

The HIIT protocol will involve 3 training sessions per week (with ≥24-48 h recovery between sessions) for 16 weeks (a total of 48 sessions lasting 36 minutes). The program will be divided into two phases: phase 1 (adaptation to HIIT training: during the first week, the intensity will be progressively increased to ensure the safety and progressive physical conditioning of the patients); and phase 2: HIIT (see **Table 3**). The high–intensity intervals will follow the Norwegian 4 x 4-minute HIIT model ^32^: 4 bouts of 4 minutes of high intensity corresponding to 85-95% HR_max_ interspersed with 3-minute active recovery intervals 54-69% HR_max_ ^33^. This protocol has been widely applied and is safe in general and in clinical populations ^34^. Additionally, this protocol includes the training parameters proposed in a recent meta-analysis ^26^ to maximize the training effects of HIIT on CRF: long intervals, high-volume and moderate to long-term duration^26^. Finally, this HIIT protocol was developed to achieve an energy expenditure similar to the MICT protocol ^35^.

**Table 3.** High-Intensity Interval Training (HIIT) Progression.

|  | **PHASE 1**<br>**(Adaptation)** | **PHASE 2**<br>**(HIIT)** |  |  |  |
| --- | --- | --- | --- | --- | --- |
| **Training Week** | **Week 1** | **Week 2-4** | **Week 5-7** | **Week 8-10** | **Week 11-16** |
| **Weekly sessions** | 3 | 3 | 3 | 3 | 3 |
| **Volume (min)** | 36 | 36 | 36 | 36 | 36 |
| **Intensity:**<br>**High Intensity**<br>**Interval** | 70-85% HRmax<br>/5-6 RPE | 85-89% HRmax<br>/6-7 RPE | 87-91% HRmax<br>/7-8 RPE | 89-93% HRmax<br>/8-9 RPE | 91-95% HRmax<br>/8-9 RPE |
| **Low Intensity**<br>**Interval** | 54-69% HRmax<br>/3-5 RPE | 54-69% HRmax<br>/3-5 RPE | 54-69% HRmax<br>/3-5 RPE | 54-69% HRmax<br>/3-5 RPE | 54-69% HRmax<br>/3-5 RPE |

HRmax: Maximal Heart Rate; RPE: Rate of Perceived Exertion (1-10 scale).

The intensity will be controlled through RPE (1-10 scale for easier interpretation by participants) and % HR_max_; a combination shown to improve adherence compared with RPE alone ^36^. Following the Guidelines for HIIT Prescription and Monitoring in clinical populations ^37^ the steps below will be implemented to validate and adjust individualized target HR zones for HIIT and educate participants and instructors regarding the correct use of RPE: i) measure HR_max_ via ramped Bruce test; ii) calculate individual target HR zone (85-95% HR_max_); iii) validate with a 4-min interval from RPE 6–9 while monitoring HR; iv) adjust the zone if HR falls outside the target despite RPE, ensuring accurate exercise prescription.

Each session will include 3 parts: warm-up (6 min), main HIIT workout (25 min), and a cool-down (5 min). Warm-up will include mobility, core exercises, and aerobic exercise involving major muscle groups at low intensity and 3 minutes of progressive aerobic exercise up to the target HR ^36^. Further details about the warm-up and materials used during the interventions are provided in **Supplementary Tables 2 and 3**. The main workout (HIIT) will include exercises involving major muscle groups and will be performed using various machines (treadmill, rower, bike, and skiers). This will ensure that the exercises can be performed by individuals regardless of potential physical limitations (e.g., limitations in upper or lower limb function). This may also enhance enjoyment and adherence. To maximize the homogeneity of the training stimulus, the equipment order during the intervals will be kept consistent across all participants. The cool-down phase will include aerobic exercise at a lower intensity, stretching, and relaxation exercises (RPE<4).

Following the guidelines for HIIT prescription and monitoring in clinical populations ^37^, the highest HR and final RPE for each high-intensity interval will be registered to achieve the four following exercise training intensity indicators for each session: 1) Peak training HR (i.e., highest HR during the entire HIIT session); 2) Average training HR (i.e., averaging highest HR from each interval); 3) Peak training RPE (i.e., highest RPE during the entire HIIT session), and 4) Average training RPE (i.e., averaging final RPE from each interval).

#### Group 2: MICT intervention program

The MICT protocol will involve 3 training sessions per week (with ≥24-48 h recovery between sessions) for 16 weeks (a total of 48 sessions lasting 44 minutes). After the first week of adaptation to the program (RPE 3-4, 54-69%HR_max_), the workout will be performed at RPE 4 to 6 (69-76%HR_max_) (see **Table 4**) to elicit an energy expenditure comparable to that of the HIIT group ^35^.

**Table 4.** Moderate-Intensity Continuous Training (MICT) Progression.

|  | **PHASE 1**<br>**(Adaptation)** | **PHASE 2**<br>**(MICT)** |  |
| --- | --- | --- | --- |
| **Training week** | 1 | 2-7 | 8-14 |
| **Weekly sessions** | 3 | 3 | 3 |
| **Volume (min)** | 44 | 44 | 44 |
| **Intensity** | 54-69%HR<sub>max</sub> /<br>3-5 RPE | 69-72%HR<sub>max</sub> /<br>3-5 RPE | 72-76%HR<sub>max</sub> /<br>3-5 RPE |

HR<sub>max</sub>: Maximal Heart Rate; RPE: Rate of Perceived Exertion (1-10 scale).

Each session will include 3 parts: warm-up (5 min), main MICT workout (34 min), and a cool-down (5 min). The warm-up will follow the same structure described previously, including mobility, core and aerobic exercise involving major muscle groups (**see supplementary table 3**), and using the same materials and rationale as indicated for the HIIT group. The cool-down phase will include aerobic exercise at a lower intensity, stretching and relaxation exercises (RPE<4). The highest and average HR and RPE for each session will be registered.

HIIT and MICT sessions will be held in small groups of up to five participants, allowing concurrent training with individualized supervision. Sessions will take place in a quiet room at HUVN or HUT, Granada (Spain), and will be supervised by exercise professionals holding a degree in Physical Activity and Sport Sciences, with specialized training in exercise for the prevention of cardiovascular diseases, along with at least one member of the study’s medical team.

#### Group 3: Control Group

The control group will follow the usual care regimen during the intervention period and will not participate in the supervised exercise sessions. This typically includes medical check-ups, with adjustments to medications or requests for any tests deemed necessary, along with encouragement to maintain a healthy lifestyle. Participants randomized to this group will be offered the opportunity to join a similar intervention after the study is completed (waiting-list control group).

#### Participant Retention and Adherence

Adherence will be tracked daily by the trainer using a registration form, recording attendance, punctuality, adverse events, and participant engagement. Sessions will be offered at multiple morning and afternoon times to facilitate attendance. Missed sessions will be followed up and rescheduled. To maximize adherence, strategies such as music during sessions, weekly motivational messages, and monthly videos via WhatsApp will be used.

### Patient and Public Involvement

Patients with FH play a key role in multiple stages of the UPPA-FH study: 1) Conception and hypothesis generation: During medical consultations, patients shared concerns about the safety and efficacy of high-intensity exercise, shaping research questions and hypotheses. 2) Study promotion and recruitment: Participants will help publicize the trial within their networks, enhancing enrollment. 3) Intervention refinement: Patients will provide real-time feedback on HIIT and MICT protocols, allowing adjustments to session pacing, rest intervals, and target HR zones to improve tolerability and adherence. They will also co-design educational materials, such as videos and brochures, to maximize clarity and relevance for the FH community. Study results will be shared via hospital newsletters, webinars with patient organizations, social media infographics, and press releases to reach the general public.

### Patient Safety

#### Criteria for Interrupting Study Participation

Participant withdrawal: Any participant may leave the study at any time without explanation or impact on their medical care.

Researcher-initiated discontinuation: The research team may withdraw a participant if continued involvement poses safety risks or violates the study protocol, including: i) severe skeletal muscle injury that alters one’s normal lifestyle (as a consequence, or not, of participating in the study); ii) serious illness preventing exercise; iii) a change of residence preventing follow-up assessments; iii) death.

#### Adverse Events

The adverse events, during the entire study period, will be systematically registered. Death, life-threatening events (e.g., stroke, myocardial infarction, fracture, etc.), or hospital admissions will be considered serious adverse events. Intervention-related dizziness, light-headedness, back or shoulder pain, or muscle aches during the study period will be considered minor adverse events.

### Potential risks, Adverse Events, and Contingency Plan

#### Risk 1: Possible development of exercise-induced myopathy in patients taking statins

Contingency plan: High-intensity exercise is not contraindicated for patients on statins, but precautions are needed to minimize myopathy risk ^38^. Before the intervention, patients will be screened for hypothyroidism (TSH), vitamin D deficiency, and concomitant myopathy-risk medications, which will be corrected or adjusted as needed. Exercise will be prescribed with gradual, individualized progression, avoiding eccentric movements when possible. Muscle pain will be assessed at each session, and creatine-phosphokinase levels measured if myalgias are intense or persistent.

#### Risk 2: Potential cardiovascular complications during the exercise program

Contingency plan: Although HIIT is generally safe, even in high-risk groups ^39^, this study will apply extra precautions to maximize participant safety. Eligibility will be limited to individuals without clinical CVD and, for those >45 years, with a normal coronary CT angiogram within 24 months. All participants will complete a ramped Bruce exercise test to confirm hemodynamic stability and exercise tolerance. HIIT volume, intensity, and progression will be individualized based on fitness and test results. Sessions will be supervised by sports science specialists and internal medicine physicians in a hospital setting to ensure immediate medical support.

#### Risk 3: Radiation Protection and Safety of Radiation Sources

Contingency plan: For PET/CT, 18F-FDG doses and CT parameters will be weight-adjusted and optimized following IAEA ^40^ and ICRP ^41^ recommendations and ALARA (As Low As Reasonably Achievable) principle. Typical 18F-FDG doses range 0.08–0.13 mCi/kg, with this study using a digital scanner allowing ∼0.07 mCi/kg. CT protocols are patient-specific and low-dose, and total exposure will be documented. After injection of the radiopharmaceutical, the patient remains at rest for approximately 2 hours in a quiet room before undergoing the scan. Once the examination is completed standard safety measures are followed: hydration, urinate frequently, and avoid close contact with pregnant women or children for 12–24 hours. Bone densitometry (DXA) uses very low radiation (0.001–0.01 mSv) and standard radiological protection principles are followed. Companions are not allowed during the test, but no post-test restrictions apply.

#### Risk 4: Low adherence to the program

Contingency plan: HIIT was chosen for its higher adherence compared to other exercise modalities ^24^. To further improve adherence, sessions will use varied equipment and music, be held in small groups (≤5 participants) for individualized guidance, and offered in morning or afternoon slots. Missed sessions will trigger proactive follow-up calls to reschedule and ensure completion.

### Statistical Analysis

Descriptive statistics will be presented as mean (standard deviation) or median (interquartile range) for quantitative variables, and as frequency and percentage for qualitative variables. In the cross-sectional study, the association of physical activity and CRF with cardiometabolic outcomes—including markers of subclinical atherosclerosis (vascular inflammation, arterial stiffness, carotid intima-media thickness and plaque) and the metabolomic profile—will be examined using correlation analyses and linear regression models. Moderation and mediation analyses will also be performed where appropriate, and the model assumptions (e.g. normality, linearity and homoscedasticity) will be assessed. Given the hypothesis-generating nature of the cross-sectional analyses, the main analyses will not be corrected for multiple testing. However, the metabolomics-related outcomes will be adjusted for false discovery rate as required.

For the RCT, effects on the primary outcome (CRF) and secondary outcomes will be analyzed on the intention to treat principle with linear mixed-effects models including fixed effects for group, time, and the group × time interaction, with a random intercept for participants. Missing data will be handled under the missing-at-random assumption using maximum likelihood within mixed models or, when necessary, multiple imputation. If evidence suggests a mechanism other than missing-at-random, an alternative appropriate method will be applied and documented. Sensitivity (i.e, per-protocol) analyses will be applied to assess the robustness of the results. Per-protocol analyses will include only participants who complete the protocol, with participants in the exercise-group required to attend ≥75% of prescribed sessions. Metabolomic analyses will be conducted using complete-case analysis. All analyses will be performed by researchers blinded to group allocation using Stata v.16 or higher (StataCorp LP, Texas, USA) and/or R, with two-sided statistical significance set at α=0.05.

### Data management and governance

All data will be managed in compliance with institutional policies, the GDPR (EU 2016/679), and Spanish Law 3/2018, guaranteeing participants’ rights. The study has ethical approval from CEI/CEIM Granada (ref. 1417-N-23). At enrolment, participants will be assigned anonymized alphanumeric codes, with personal identifiers stored separately in an encrypted, access-restricted file. Research data will be pseudonymized and stored on a secure, password-protected institutional drive accessible only to authorized investigators.

A data monitoring committee will not be required given the study design, sample size, the team’s data-management expertise, and the non-pharmacological nature of the trial. Data will be periodically reviewed for completeness and consistency, with regular backups to ensure integrity. At study completion, pseudonymized data will be retained for at least 10 years before secure anonymization and archiving.

Consent forms and essential documents will follow Good Clinical Practice and Spanish biomedical regulations. Data management will comply with FAIR principles (Findable, Accessible, Interoperable and Reusable), ensuring standardized metadata, open formats, and transparent documentation. In line with open-science practices, anonymized datasets may be shared upon reasonable request after publication.

### Potential Impact of the UPPA-FH Study

Familial hypercholesterolemia remains underdiagnosed and undertreated, with many patients failing to reach LDL-c targets despite intensive therapy, highlighting the need for complementary strategies. Exercise has well-documented cardiometabolic benefits, making its evaluation in FH timely and clinically relevant. The UPPA-FH trial is the first RCT on exercise in FH and comparing HIIT and MICT on CRF and subclinical atherosclerosis. By integrating metabolomics and assessing safety, the study will clarify both mechanistic pathways and the feasibility of structured exercise in this high-risk population.

If successful, findings may support supervised exercise as a scalable, non-pharmacological adjunct to standard therapy. Strengths include the RCT design and objective endpoints (accelerometry, imaging, metabolomics), though the 16-week, 18– 70-year-old, cardiovascular-only intervention may limit long-term and external validity. Future studies should explore longer, multi-modal programs in more diverse populations and assess the sustainability of effects.

### Dissemination Policy

Findings will be shared through publications in peer-reviewed scientific journals and presentations at international conferences. The primary objective is to publish a main manuscript on the primary outcome, with additional articles addressing secondary outcomes. The team will also engage the public through the FH Foundation, patient associations, press releases, guides for primary care, hospital newsletters, webinars, and social media infographics.

## Data Availability

Data management will comply with FAIR principles (Findable, Accessible, Interoper-able and Reusable), ensuring standardized metadata, open formats, and transparent documentation. In line with open-science practices, anonymized datasets may be shared upon reasonable request after publication.

## Acknowledgments

We thank all participants for their contributions and the staff of Internal Medicine, Cardiology, Nuclear Medicine, Radiology, and Clinical Analysis at HUVN for supporting the study.

## Author’s contribution

All authors made substantial intellectual contributions to the conception and design of the protocol of this study. JAV-H, PG-B, BG-C and AS-M designed the study. The first draft was written by JAV-H, with important revisions and contributions from AS-M and BG-C. AS-M and JAV-H served as principal investigators, project administrators, and guarantors, and they assume full responsibility for the completed work and the conduct of the study. All authors have reviewed and approved the final manuscript, given written consent for its publication, and accept full accountability for its contents. The contributions of each author were agreed upon prior to manuscript submission.

## Patient and public involvement

Patients and/or the public were involved in the design, conduct, reporting, or dissemination plans of this research. Refer to the Methods section for further details.

## Data sharing

Data management will comply with FAIR principles (Findable, Accessible, Interoperable and Reusable), ensuring standardized metadata, open formats, and transparent documentation. In line with open-science practices, anonymized datasets may be shared upon reasonable request after publication.

