## Supplementary Material for "Unraveling the Potential of Physical Activity and High-Intensity Interval Training in the Treatment of Familial Hypercholesterolemia: Impact on Cardiorespiratory Fitness, Atherosclerosis, and Underlying Mechanisms. Study protocol of the UPPA-FH Project"

**Supplementary Material for the Study Protocol of the UPPA-FH Project**

**Methods**

**A. Description of Secondary Outcomes**

**1. Subclinical atherosclerosis**

*1.1. Vascular Inflammation*

Evaluated by performing a PET/CT. This examination will be performed with a multimodal PET/CT scanner with LSO crystal detectors (Siemens Biograph Vision 600, from Siemens Healthcare®, Erlangen, Germany). The protocol for the preparation procedure, administration of the radiopharmaceutical, and image acquisition will be based on the available international recommendations (1,2).

Patients will be instructed to follow a low-carbohydrate, high-protein, high-fat diet for at least 24 hours before 18F-FDG PET/CT in order to suppress physiological myocardial uptake, and they will avoid strenuous physical activity on the day before imaging. They will fast for a minimum of 4 hours before their appointment, remain well hydrated, and refrain from consuming sugary beverages or stimulants. Upon arrival, capillary blood glucose will be measured and, if it exceeds 200 mg/dL, the study will be deferred and rescheduled. 18F-FDG (2-fluoro-2-deoxy-D-glucose) will be administered at a dose of 10 MBq per kilogram of body weight and injected intravenously through a peripheral venous catheter, with the line flushed using 5–10 mL of normal saline before and after injection to ensure complete delivery and maintain catheter patency. Following radiotracer administration, patients will rest supine in a quiet uptake room for 120 minutes; they will be asked to void immediately before scanning and to remove all metallic objects. Imaging will then be performed according to our institution’s standard PET/CT protocol, in two sequential phases. First, a whole-body scan will be acquired from the supraclavicular region through the pelvis (with patients’ arms elevated above the head), using 6 minutes per bed position to evaluate the thoracic and abdominal aorta as well as the iliac arteries. Second, a dedicated head and neck acquisition will be performed with the head secured in an immobilizer and the arms resting alongside the body: a single 8-minute bed position encompassing the skull and cervical regions to assess the carotid arteries. Image interpretation will focus on the evaluation of arterial-wall inflammation across multiple vascular territories. Qualitative assessment of FDG uptake will be performed visually, while quantitative analysis will employ the target-to-background ratio (TBR); a vessel will be considered inflamed when the TBR will be ≥1.6. The TBR will be defined as the ratio of the maximum standardized uptake value (SUVmax) within atherosclerotic plaques or the arterial wall to the mean SUV of the venous blood pool, thus correcting for circulating tracer activity. This metric will correlate closely with plaque macrophage density on corresponding histological sections. TBR measurements will be obtained for carotid arteries, thoracic aorta, abdominal aorta, and iliac arteries (3).

*1.2. Arterial Stiffness*

Evaluated by determining the pulse wave velocity (PWV), with higher velocities indicating greater stiffness. Its elevation is an early marker of atherosclerosis. The Mobil-O-Graph® 24h pulse wave monitor device (IEM GmbH, Stolberg, Germany) will be used, whose operation is based on oscillometry recorded by a blood pressure cuff placed on the brachial artery. A value higher than 10 m/s will be considered pathological (4).

*1.3. Intima-Media Thickness*

Evaluated by carotid doppler ultrasound with General Electric's Logic F6® ultrasound device, considering markers of subclinical atherosclerosis both a carotid intima-media thickness (IMT) greater than 1.0 mm and/or the presence of carotid plaque, (characterized as a localized thickening of the intima-media layer reaching at least 1.5 mm, protruding into the lumen, or exceeding the surrounding IMT by at least 0.5 mm or 50%) (5).

**2. Blood and Urine parameters**

Plasma and serum samples will be obtained from participants on the second day of assessment, as described in the data collection section. Blood will be drawn into four tubes, and the following parameters will be analyzed.

*2.1. Inflammatory Markers*:* High sensitivity C-reactive protein (mg/L).

*2.2. Markers of Oxidative Stress:* Oxidized LDL (pg/ml) (Human OxLDL ELISA Kit; with a sandwich ELISA (catalog: SEA527Hu, USCN).

*2.3. Circulating markers of Endothelial Dysfunction:*

Human soluble Inter-Cellular Adhesion Molecule-1 (sICAM-1)/CD54 (pg/mL) with a commercial ELISA kit (Ray Biotech).

Homocysteine* (µmol/l; AxSYM Homocysteine; Abbott Laboratories, Abbott Park, IL).

*2.4. Muscle metabolism parameters*:* Vitamin D (nmol/L) and TSH (mUI/L).

*2.5. Glucose and lipid profile*:* Glucose (mg/dL), triglycerides (mg/dL), total cholesterol (mg/dL), LDLc (mg/dL) and HDLc (mg/dL), apoproteins and lipoprotein a.

*2.6. Coagulation*:* Fibrinogen (mg/dL) will be measured by von Clauss assay.

*2.7. Markers of renal function** as urea (mg/dL) and creatinine (mg/dL)

*2.8. Hemogram*:* Erythrocytes (x 106/uL), hemoglobin (Hb, g/dL), mean corpuscular volume (VCM, fL), leukocytes (x 103/uL) and platelets (x 103/uL).

*2.9. Microalbuminuria*:* Albumin (mg/dL) in urine by nephelometry (Hy-Pro 50 protein analyzer).

**Indicated biochemical parameters* will be determined following standardized methods at the Hospital Universitario Virgen de las Nieves or Hospital Universitario Torrecárdenas with an Alinity autoanalyzer with reagents supplied by the company Abbot ® according to the routine procedures of our hospital.

For the remaining markers, instructions provided by the corresponding manufactueres will be followed and plates will be read espectrophotometrically at the indicated wavelength with a BioTek plate reader (Agilent).

*2.10. Metabolomics:* Untargeted metabolomic profiles will be acquired by Nuclear Magnetic Resonance (NMR) using a Bruker Avance Ascend HD 600 MHz spectrometer equipped with a quadruple resonance HCNP cryoprobe and a thermostated SampleJet autosampler with capacity for up to 480 samples. Serum aliquots (300 µL) will be mixed with 300 µL of deuterated phosphate buffer (0.075 M KH_2_PO_4_ in D_2_O, pH 7.4), containing 0.1% (w/v) of the sodium salt of 3-(trimethylsilyl)propionic-2,2,3,3-*d*_4_ acid (TSP) as a chemical shift reference and 90 µM sodium azide (NaN₃) as an enzymatic inhibitor. The resulting 600 µL mixtures will be transferred into 5 mm NMR tubes for spectral acquisition. All spectra will be recorded at 300 ± 0.1 K. A Carr-Purcell-Meiboom-Gill (CPMG) pulse sequence with presaturation (Bruker 1D cpmgpr1d) will be used to attenuate broad resonances arising from macromolecules and proteins. This approach is expected to allow the detection and quantification of approximately 40 metabolites and metabolic classes, including carbohydrates (e.g., glucose), amino acids and derivatives (e.g., creatine), tricarboxylic acid (TCA) cycle intermediates (e.g., citric acid, pyruvic acid), choline-containing compounds (essential for membrane structure), nucleotides/nucleosides, polyols (e.g., glycerol), fatty acids, triacylglycerides, and ketone bodies (e.g., 3-hydroxybutyrate, acetate, and acetoacetate). In addition to CPMG, two complementary NMR experiments will be acquired. First, standard NOESY spectra with water suppression (Bruker 1D noesygppr1d) will be recorded to enable quantification of specific small-molecule resonances of interest. Subsequently, diffusion-ordered spectroscopy (Bruker 1D stegpbp1s1d) will be performed at two different diffusion times to allow precise quantification of lipoprotein subclass distributions (VLDL, LDL, IDL, and HDL), supramolecular phospholipid composite (SPC), glycoproteins A and B, and total triglycerides. Triglyceride concentrations obtained from diffusion experiments will be cross-validated with those measured by CPMG and NOESY methods. Acquiring diffusion-weighted spectra at two distinct diffusion times will also help confirm whether the measured proportions of these supramolecular assemblies reflect their actual abundance or are biased by differential transverse relaxation due to distinct molecular correlation times. All NMR data, along with additional variables, will be analysed using multivariate statistical techniques to identify relevant biomarkers and develop predictive models. These analyses will be carried out by the NMRMBC group at the University of Almería.

**3. Other Outcomes of Interest**

*3.1. Physical activity:* Assessed with triaxial accelerometers (Axivity AX3, Axivity Ltd, United Kingdom) to characterize the physical activity status of the participants both in the cross-sectional study and the RCT. The AX3 has been validated and used to measure physical activity in a previous large-scale cohort study (6). Participants will use the accelerometer on the non-dominant wrist 24 hours a day for 10 consecutive days. A sampling frequency of 25 Hz will be set. Unprocessed accelerometry data will be downloaded as ".cwa" files and processed with R software using the GGIR package (v. 2.5-0, <https://cran.r-project.org/web/packages/GGIR/>) (7). The most current thresholds will be applied to calculate the amount of time in sedentary activities, light, moderate, vigorous, and very vigorous activities.

*3.2. Anthropometric Measures and Body Composition:* Height (cm) and body weight (kg) will be measured using a Seca 799 digital column scale with integrated stadiometer (Seca GmbH & Co. KG, Hamburg, Germany; Mat. No. 7997021099). Height will be recorded via a sliding headpiece on the metal stadiometer, with a measurement range of 60–200 cm and 1 mm precision. Body mass index (BMI) will be calculated in kg/m^2^. Waist and hip circumference will be measured by non-elastic anthropometric tape (SECA 200). Bone mineral content (BMC), bone mineral density (BMD), fat mass, lean mass, and fat-free mass will be measured via DXA using a Discovery Wi system (Hologic, Bedford, MA, USA) in accordance with the manufacturer’s standard scanning protocol. (8). Participants will be positioned supine with limbs aligned alongside the body, and scans will be acquired and processed using the system’s dedicated software. Derived indices [including fat-free mass index (FFMI), lean mass index (LMI), and fat mass index (FMI)] will be calculated by normalizing each compartment’s mass (in kg) to height squared (m²). Total fat mass will be additionally expressed as a percentage of the participant’s total body weight.

*3.3. Muscular Strength and Function:* Lower-body muscular strength and function will be evaluated using the 30-second chair stand test(9). The number of times a participant could stand up from a seated position to a full upright posture, with arms crossed over the chest, will be recorded. Upper-body muscular strength will be measured with a digital handgrip dynamometer (TKK 5101 Grip-D; Takey, Tokyo, Japan). Each participant will perform two trials, alternating hands, with a one-minute rest between attempts. The best score from each hand (dominant and non-dominant) will be averaged to calculate the overall mean handgrip strength(10).

*3.4. Sociodemographic and Clinical Variables:* Age, education level, occupational status, family history of CVD, personal history of CVD, treatments, and questions regarding female participants’ health status will be recorded. In addition, 5-year and 10-year cardiovascular risk estimates will be calculated using the SAFEHEART-Risk Equation (11).

*3.5. Mediterranean Diet:* The adherence to the traditional Mediterranean Diet will be evaluated using the 14-point questionnaire from the PREDIMED trial (12), which has been validated for this purpose. The PREDIMED questionnaire consists of twelve questions about the frequency of consumption of essential foods, along with two questions concerning specific dietary habits associated with the Mediterranean Diet. Each question is scored either 0 or 1 point, with the total score ranging from 0 to 14. A score of 0 indicates no adherence, while a score of 14 represents full adherence to the Mediterranean Diet.

*3.6. Health-related Quality of Life:* The Spanish version of the SF-36 (13) will be used to evaluate Health-Related Quality of Life (QoL). The SF-36 consists of 36 items organized into 8 dimensions (physical function, physical role, bodily pain, general health, vitality, social functioning, emotional role, and mental health), along with 2 summary components (physical and mental health). The scores range from 0 to 100, where higher scores indicate better QoL.

*3.7. Anxiety and Depression:* The presence of anxiety and depression in patients will be assessed with the Hospital Anxiety and Depression Scale (HADS). It consists of 14 items, each with four possible responses (ranging from 0 to 3 points), divided into two subscales: seven items assess depression, and the remaining items assess anxiety. The maximum score for each subscale is 21 points, with scores below 11 indicating the presence of depression or anxiety (14).

**B. Supplementary Tables**

**Supplementary Table 1**. Ramped Bruce protocol (15).

| **Stage** | **Time** | **Speed (Km/h)** | **Grade (%)** | **METS (aprox)** |
| --- | --- | --- | --- | --- |
| Start | 00:00 | 1.0 | - | - |
| Stage 1 | 00:15 | 1.6 | 0.0 | 1.4 |
| Stage 2 | 00:30 | 1.6 | 1.0 | 1.6 |
| Stage 3 | 00:45 | 1.6 | 2.0 | 1.7 |
| Stage 4 | 01:00 | 1.7 | 3.0 | 1.8 |
| Stage 5 | 01:15 | 1.9 | 4.0 | 1.9 |
| Stage 6 | 01:30 | 1.9 | 5.0 | 2.0 |
| Stage 7 | 01:45 | 1.9 | 6.0 | 2.1 |
| Stage 8 | 02:00 | 2.0 | 7.0 | 2.2 |
| Stage 9 | 02:15 | 2.2 | 8.0 | 2.3 |
| Stage 10 | 02:30 | 2.4 | 9.0 | 2.4 |
| Stage 11 | 02:45 | 2.5 | 10.0 | 2.5 |
| Stage 12 | 03:00 | 2.7 | 10.0 | 2.8 |
| Stage 13 | 03:15 | 2.8 | 10.0 | 3.0 |
| Stage 14 | 03:30 | 2.9 | 11.0 | 3.3 |
| Stage 15 | 03:45 | 3.0 | 11.0 | 3.5 |
| Stage 16 | 04:00 | 3.1 | 11.0 | 3.9 |
| Stage 17 | 04:15 | 3.2 | 11.0 | 4.0 |
| Stage 18 | 04:30 | 3.2 | 11.0 | 4.3 |
| Stage19 | 04:45 | 3.3 | 12.0 | 4.5 |
| Stage 20 | 05:00 | 3.5 | 12.0 | 4.9 |
| Stage 21 | 05:15 | 3.7 | 12.0 | 5.0 |
| Stage 22 | 05:30 | 3.9 | 12.0 | 5.4 |
| Stage 23 | 05:45 | 4.0 | 12.0 | 5.6 |
| Stage 24 | 06:00 | 4.0 | 12.0 | 5.9 |
| Stage 25 | 06:15 | 4.0 | 13.0 | 6.0 |
| Stage 26 | 06:30 | 4.1 | 13.0 | 6.3 |
| Stage 27 | 06:45 | 4.2 | 13.0 | 6.4 |
| Stage 28 | 07:00 | 4.3 | 13.0 | 6.7 |
| Stage 29 | 07:15 | 4.5 | 14.0 | 6.8 |
| Stage 30 | 07:30 | 4.7 | 14.0 | 7.2 |
| Stage 31 | 07:45 | 4.8 | 14.0 | 7.4 |
| Stage 32 | 08:00 | 5.0 | 14.0 | 7.8 |
| Stage 33 | 08:15 | 5.1 | 14.0 | 7.9 |
| Stage 34 | 08:30 | 5.3 | 14.0 | 8.3 |
| Stage 35 | 08:45 | 5.4 | 14.0 | 8.4 |
| Stage 36 | 09:00 | 5.5 | 14.0 | 8.8 |
| Stage 37 | 09:15 | 5.6 | 14.0 | 9.0 |
| Stage 38 | 09:30 | 5.6 | 14.0 | 9.3 |
| Stage 39 | 09:45 | 5.7 | 15.0 | 9.5 |
| Stage 40 | 10:00 | 5.8 | 15.0 | 9.9 |
| Stage 41 | 10:15 | 5.9 | 15.0 | 10.1 |
| Stage 42 | 10:30 | 6.1 | 15.0 | 10.1 |
| Stage 43 | 10:45 | 6.2 | 16.0 | 10.4 |
| Stage 44 | 11:00 | 6.4 | 16.0 | 10.4 |
| Stage 45 | 11:15 | 6.4 | 16.0 | 11.1 |
| Stage 46 | 11:30 | 6.5 | 16.0 | 11.1 |
| Stage 47 | 11:45 | 6.7 | 16.0 | 11.4 |
| Stage 48 | 12:00 | 6.7 | 16.0 | 11.7 |
| Stage 49 | 12:15 | 6.7 | 16.0 | 12.5 |
| Stage 50 | 12:30 | 6.9 | 16.0 | 12.8 |
| Stage 51 | 12:45 | 6.9 | 17.0 | 12.8 |
| Stage 52 | 13:00 | 7.0 | 17.0 | 13.1 |
| Stage 53 | 13:15 | 7.0 | 17.0 | 13.4 |
| Stage 54 | 13:30 | 7.2 | 17.0 | 13.4 |
| Stage 55 | 13:45 | 7.4 | 18.0 | 13.4 |
| Stage 56 | 14:00 | 7.5 | 18.0 | 13.7 |
| Stage 57 | 14:15 | 7.7 | 18.0 | 14.3 |
| Stage 58 | 14:30 | 7.8 | 18.0 | 14.3 |
| Stage 59 | 14:45 | 8.0 | 18.0 | 14.6 |
| Stage 60 | 15:00 | 8.0 | 18.0 | 14.6 |

**Supplementary Table 2:** Exercise equipment for the interventions of the UPPA-FH study

| Equipment (Brand) | Number | Characteristics | Photo |
| --- | --- | --- | --- |
| Fitness mat  (Tunturi) | 8 | Colour: Black | 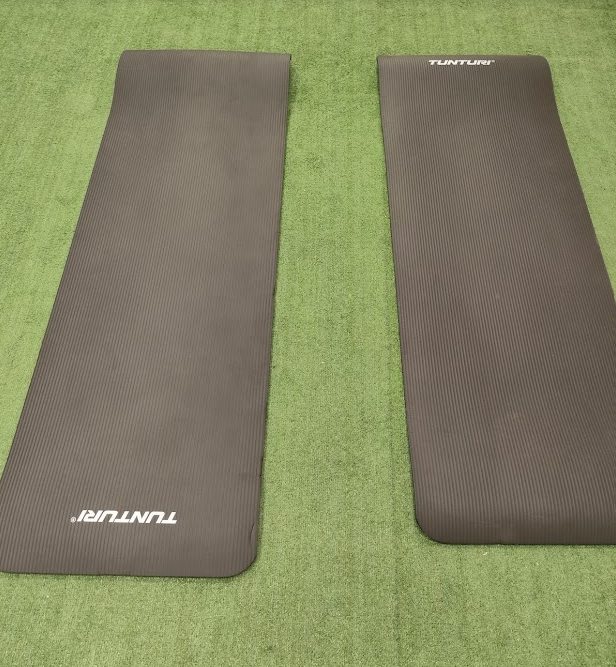 |
| Resistance Bands  (Tunturi) | 12 | Low and medium resistance | 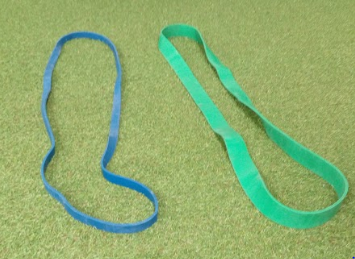 |
| Bank  (Bowflex) | 1 | Colour: Black.  Positions: 3 inclinations. | 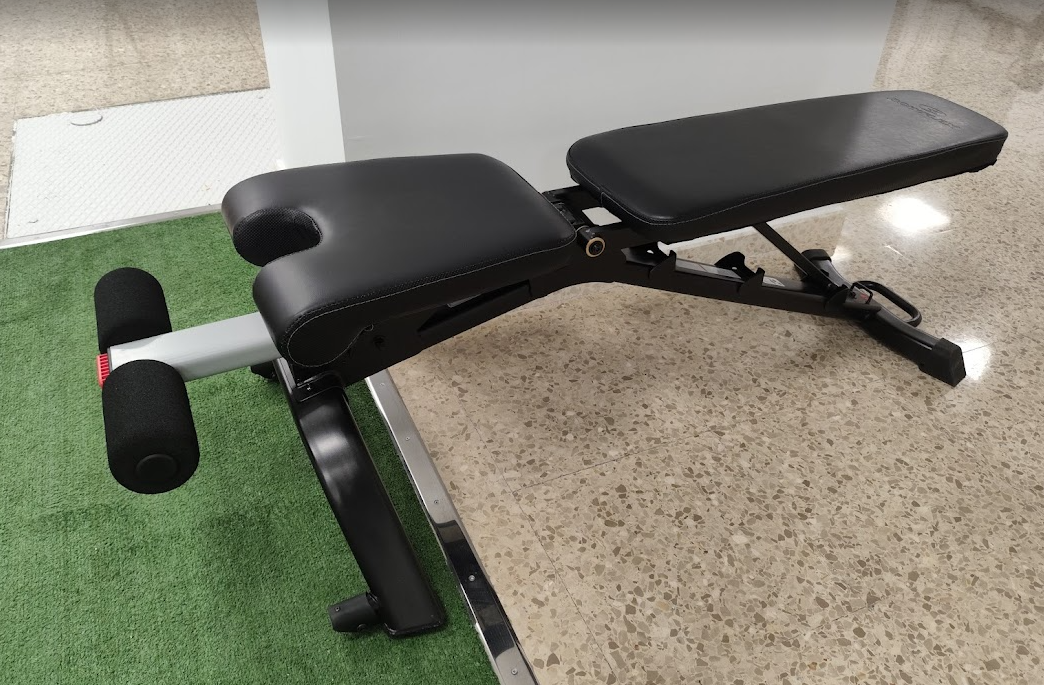 |
| Kettlebells  (Tunturi) | 3 | Colour: Black  Weight: 8/ 10/ 12 kg | 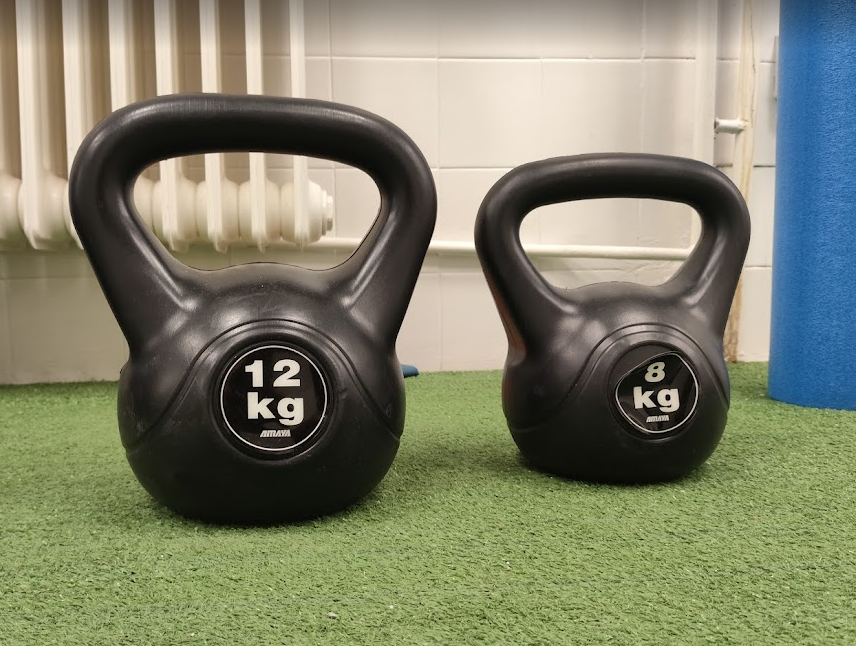 |
| Adjustable Dumbbells  (Bowflex) | 1 pair | Colour: Black.  Weight: 2 to 24 kg | 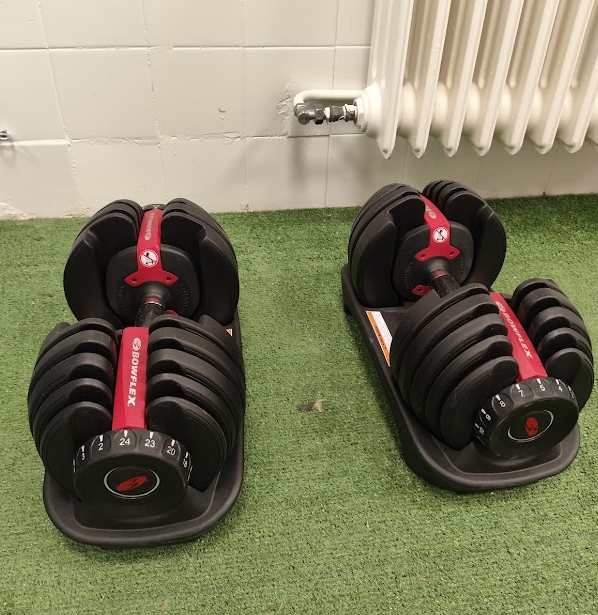 |
| Treadmill  (BH) | 5 | Model: Serie i.RC12 Dual | 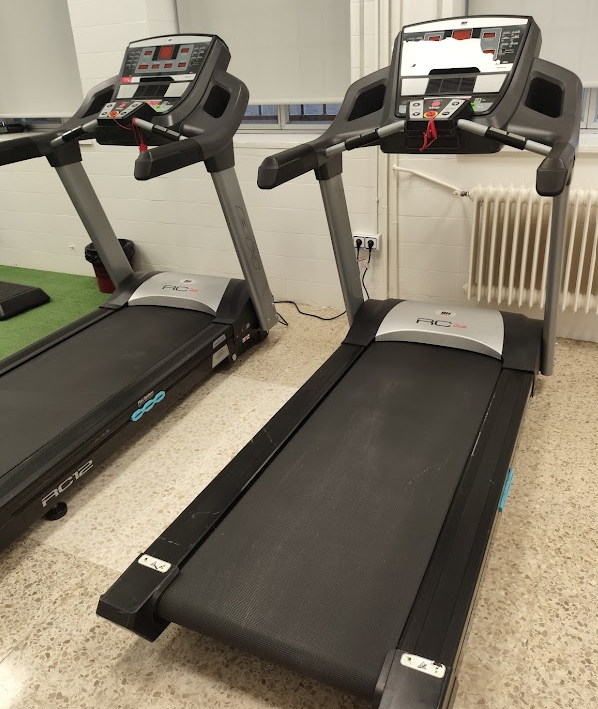 |
| Elliptical  (Reebok) | 1 | Model: SL 8.0 | 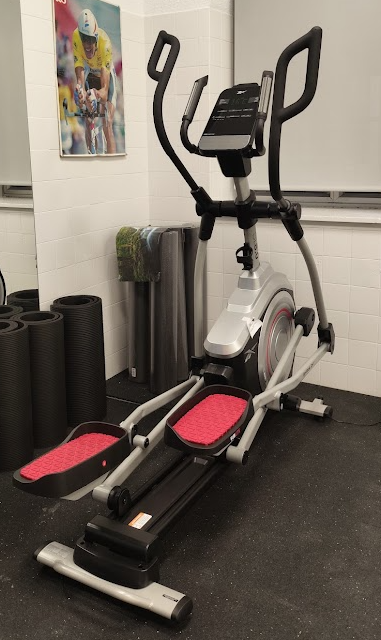 |
| Stationary bike  (Bodytone) | 1 | Model: EX25 | 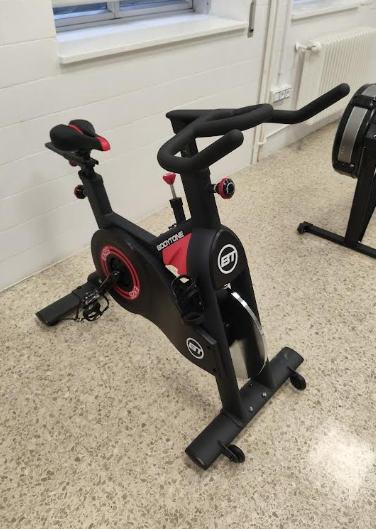 |
| Stationary bike (Concept2) | 2 | Model: BikeErg PM5 | 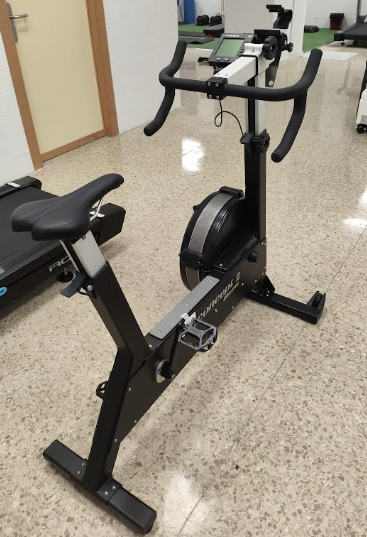 |
| Row ergometer (Concept2) | 2 | Model: RowErg PM5 | 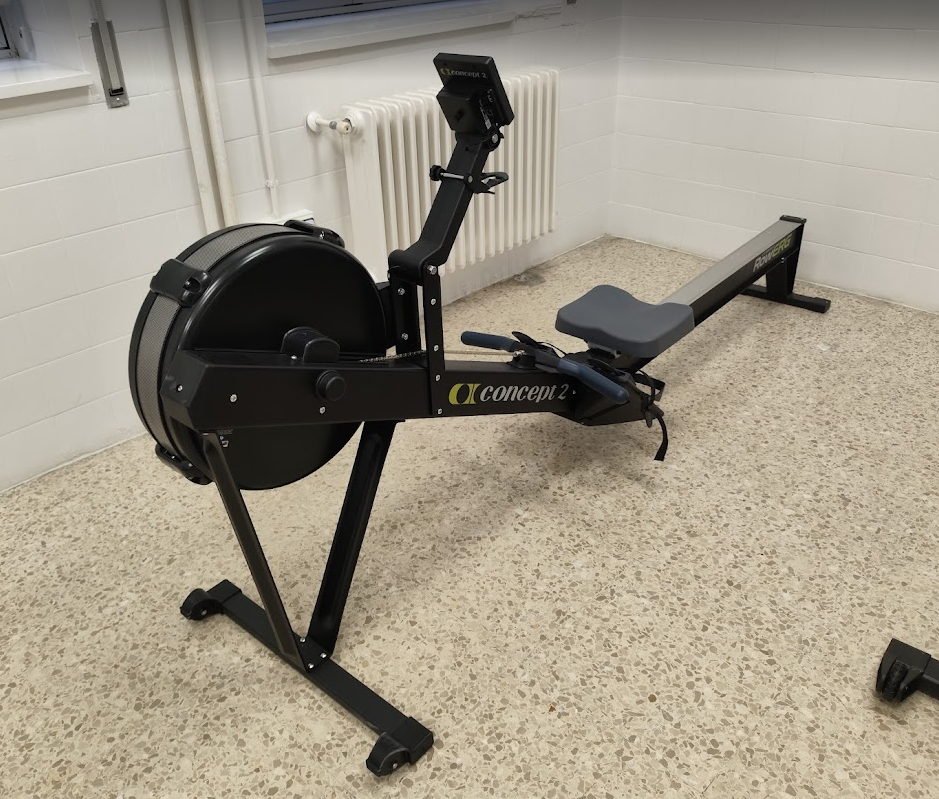 |
| Ski ergometer (Concept2) | 1 | Model: SkiErg PM5 | 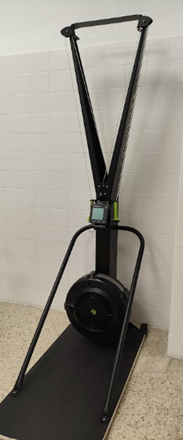 |

**Supplementary Table 3.** Detailed description of the UPPA-FH warm-up, including mobility, core, and aerobic exercise involving major muscle groups at low intensity.

**Cat-camel** | Duration: 10 seconds | Intensity: RPE 3/10

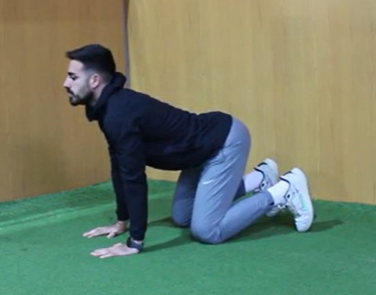

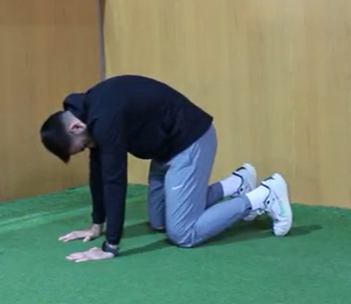

**Quadruped Rock Back** | Duration: 10 seconds | Intensity: RPE 3/10

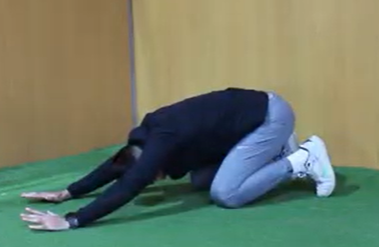

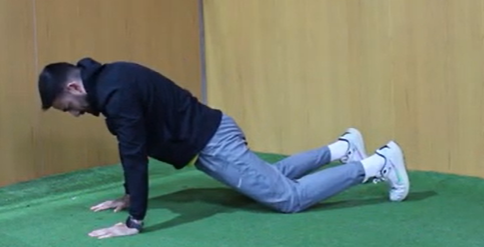

**Bird Dog** | Duration: 20 seconds | RPE: 4-6/10

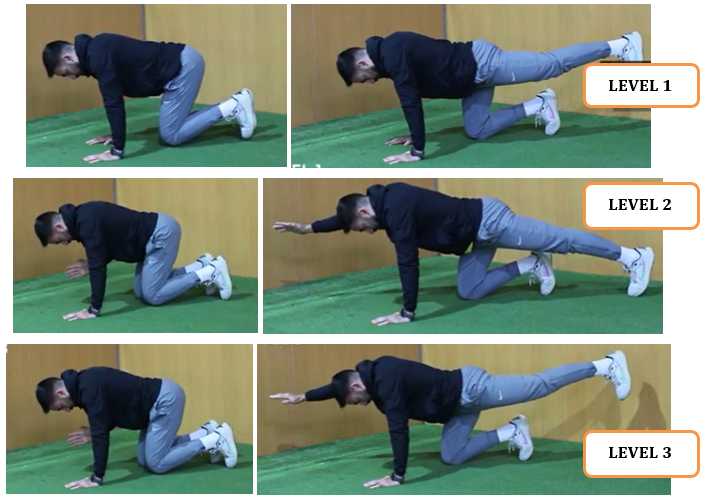

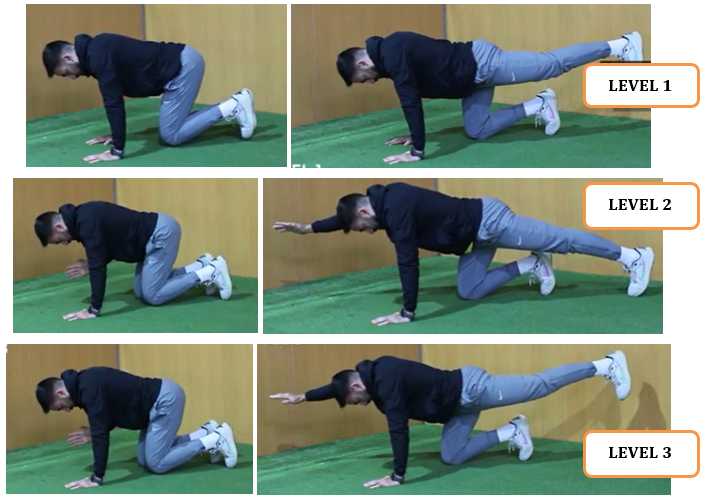

**Plank** | Duration: 20 seconds | RPE: 4-6/10

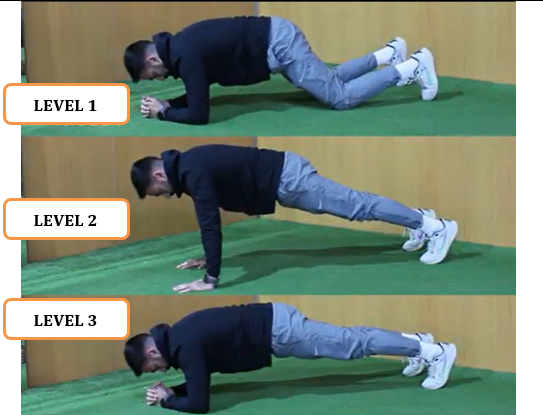

**Dead bug |** Duration: 20” | Intensity: RPE 4-6/10

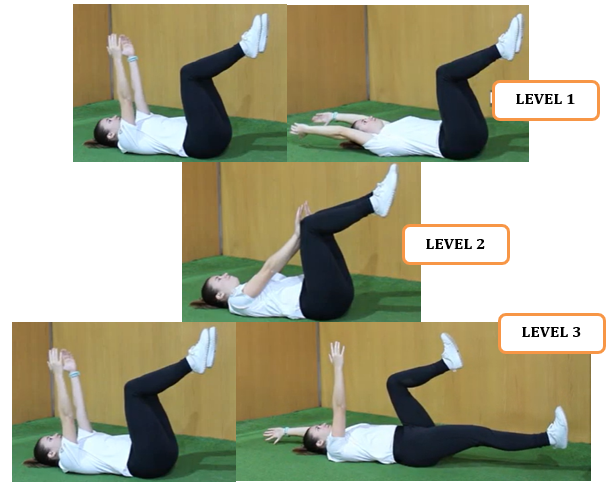

**Squat** | Duration: 20 seconds | Intensity: RPE 4-6/10

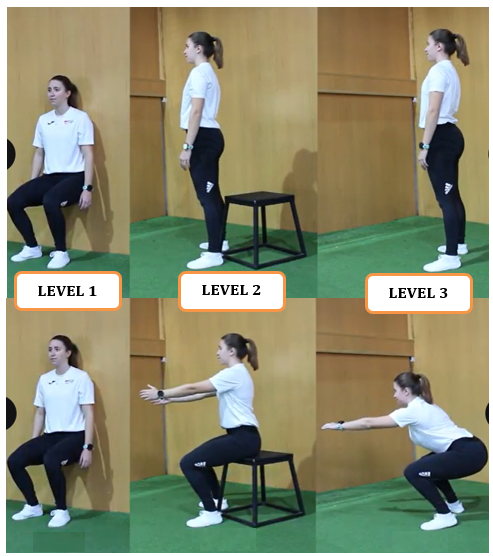

**Deadlift – hip hinge** | Duration: 20 seconds | Intensity: RPE 4-6/10

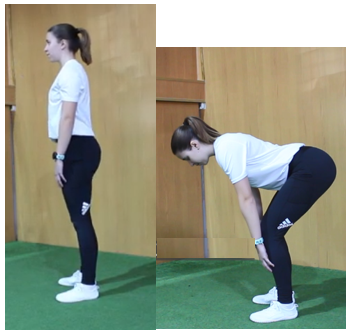

**Horizontal pull** | Duration: 20 seconds | Intensity: RPE 4-6/10

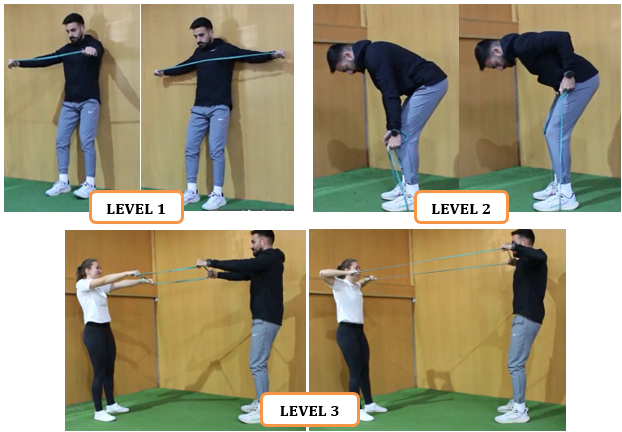

**Progressive aerobic exercise up to the target heart rate: 2-3 minutes**

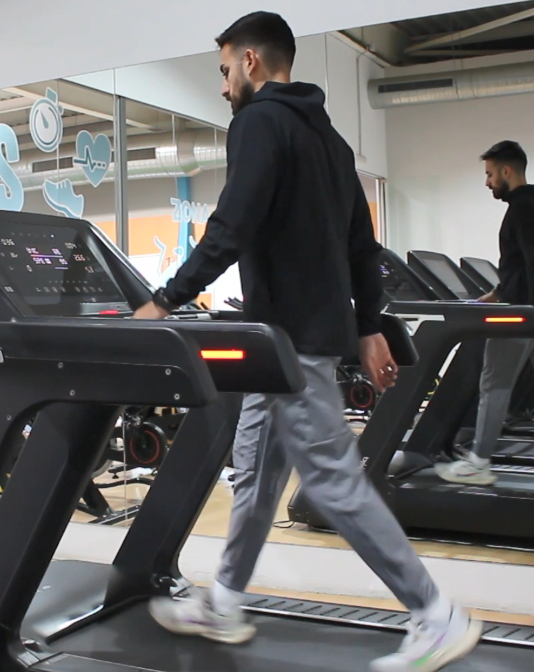

Statement of consent: All individuals depicted in the photographs are either authors of this manuscript or members of the research group and have provided their explicit consent for the inclusion and publication of their images in this preprint.
